# Know before you go! A field survey of the preparedness of wilderness day hikers and trail runners in Rocky Mountain National Park

**DOI:** 10.1101/2025.09.23.25336486

**Authors:** John T. Lambert, Davidson H. Hamer, Taylor N. Weckstein, Gregory A. Wellenius

## Abstract

**Introduction:** Actively recreating in natural environments enhances physical and mental health, but also carries risk. We aimed to characterize wilderness day hikers and trail runners, and examine factors that predict preparedness.

**Methods:** We conducted a cross-sectional survey in 4 distinct areas of Rocky Mountain National Park (RMNP) in Colorado during June-August of 2024. English-speaking adults returning from a day hike or trail run were invited to participate. We classified visitors as ‘prepared’ based on the gear they reported carrying.

**Results:** Of 801 potential participants approached, 586 (82.3%) day hikers and 68 (76.4%) trail runners agreed to participate. The overall average age was 40.7 years (range 18-82); 50.1% were female; and most common state of residence was Colorado (47.3%). Day hikers tended to be older, travel in larger groups, and spend fewer days in the wilderness per year while trail runners reported higher levels of experience and wilderness preparedness, and were more likely to experience ‘close calls’. A minority of participants met our definition of wilderness prepared, about half were altitude prepared, and approximately a quarter did not tell anyone where they were going and when they expected to return. Several measures of experience were associated with preparedness.

**Conclusions:** Both equipment and knowledge are important for safely enjoying and leaving wilderness settings. Yet many wilderness users in RMNP did not meet our definition of adequate preparation, especially those with less experience. Additional efforts to increase the proportion of wilderness day-users who are prepared may help further improve visitor safety.

## Introduction

The popularity of outdoor activities, such as hiking and off-road running, has increased dramatically in recent years, and participating in outdoor recreation increased during the COVID-19 pandemic ^1–3^. Rocky Mountain National Park (RMNP), a large wilderness park in Colorado, is one of the most visited national parks in the United States ^4,5^. RMNP encompasses 415 square miles (1075.8 sq km), 94.8% of which is designated as wilderness. The elevation in RMNP ranges from 7,600 ft to 14,259 ft (2,316-4,346 m), and includes 355 miles (571 km) of established trails ^6^. While the vast majority of visitors recreate without incident, between 2009-2011 the park averaged 229 rescue operations per year ^7^. Day hikers make up the overwhelming majority of Search and Rescue (SAR) operations in RMNP ^8^.

Many common mishaps in wilderness settings are easily preventable. For example, acute mountain sickness (AMS), dehydration, and respiratory distress can be prevented with acclimatization, access to proper hydration, and prophylaxis with medications such as acetazolamide and dexamethasone ^9,10^. When injury or illness occurs in wilderness settings, preparedness may help mitigate morbidity and mortality. However, preparedness requires knowledge and gear; carrying the ’10 essentials’ – key items that wilderness users are recommended to carry in case of problems - has been used as a proxy for studying preparedness^11^. Many visitors to RMNP, especially those visiting Longs Peak are unaccustomed to the extreme altitude ^12^. Previous research has also documented that most mountain climbers of the highest peaks in Colorado have low preparedness, often lacking medical knowledge and/or safety gear ^13^.

While the epidemiology of preparedness in some outdoor populations, such as backpackers and climbers, has been studied in both RMNP and in general, there is a dearth of peer-reviewed studies on US off-road runners ^13–15^. Off-road running includes trail runners, mountain running, skyrunning, fell running, orienteering, cross country running, and ultramarathon running (collectively referred to as ‘trail runners’) ^16^. While trail running has many benefits, it also carries a high risk of injuries ^17,18^. Trail running is semi- to fully self-sufficient, in which runners must carry their own gear, navigation, hydration, and food ^16^. Compared to hikers, trail runners often travel further distances while carrying lighter packs with minimal hydration due to weight or comfort concerns, a combination that may put them both at higher risk of injury and at higher risk of adverse outcomes if an incident occurs ^15,19,20^. Previous research has primarily focused on trail running in race environments ^21,22^. We were unable to find any previous studies specifically examining recreational trail runner preparedness and habits.

Additionally, no study has examined preparedness among visitors in RMNP following the COVID-19 pandemic (post-2020), a period when park usage first dropped significantly, and subsequently surged ^4^. During the COVID-19 pandemic, after initial stay-at-home orders ended, many people turned to outdoor recreation as a safe, healthy, and beneficial activity - a substantial portion of whom were new or inexperienced ^3,23^.

To address these knowledge gaps, we conducted a cross-sectional survey in RMNP in 2024 during the three busiest months of the year. Our goals were to: 1) characterize the demographics, experience, and preparedness of post-pandemic wilderness day users and 2) examine the association between experience and preparedness among both trail runners and day hikers.

## Methods

We administered a cross-sectional survey to a convenience sample of day hikers and trail runners on backcountry trails in RMNP in June, July, and August 2024. RMNP was chosen because of its popularity, varied elevation, and generalizability to other large wilderness parks. More than 4 million visitors enter RMNP each year, and more than half of the total visits each year occur during these 3 months ^4^. We approached day hikers and trail runners in 4 areas of the park (Figure 1; Bear Lake, Longs Peak, Wild Basin, Colorado River District). Each area of the park was sampled on a rotating basis on both weekends and weekdays throughout the 3 months, for a total of 38 sampling days. These areas were selected to provide a sample of users across trails that varied in characteristics, popularity, and difficulty level. Longs Peak was of particular interest as it has high use, high altitude, most of the trail is above tree line and therefore very exposed, with frequent extreme weather ^24^. The Bear Lake area is one of the most visited locations in the park, and is often recommended for its network of trails, accessibility, and lakes ^25,26^. The other trailheads offer hikes of varying difficulty, popularity, terrain, elevation, and other attributes. National Park Service (NPS) signage at the beginning of all trailheads surveyed listed essential items to carry, the hazards of high altitude (including the symptoms of AMS), a reminder to let someone know where you’re going and when to expect you back, regulations (including stay on trail), a map, hazard information, and more (supplemental figure 1).

**Figure 1:**
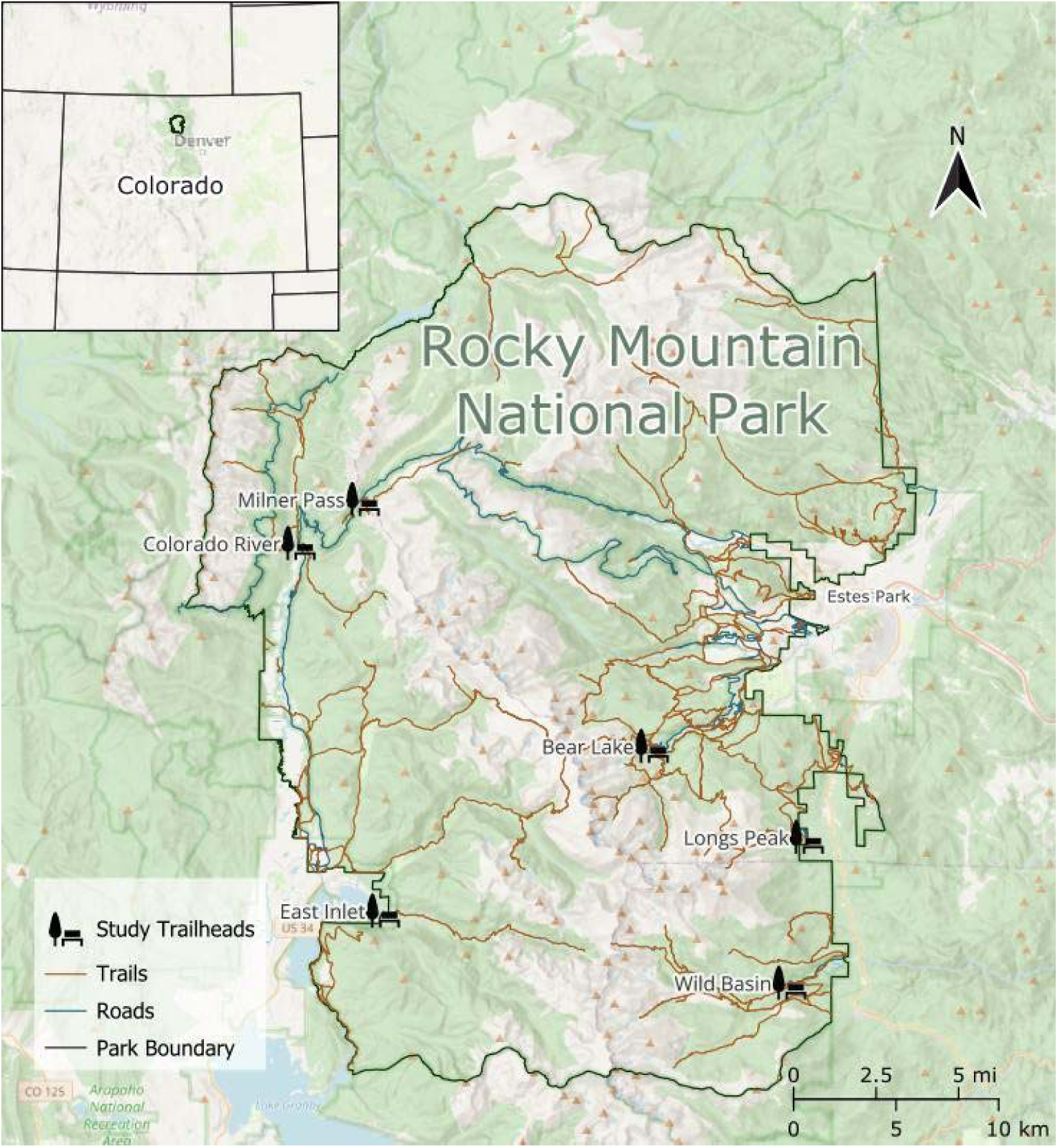
Study Trailheads. A map of the park showing the trailhead locations that were surveyed within RMNP. Base map data: Copyright OpenStreetMap, data: National Park Service.

We approached potential participants as they were returning from their hike/run to minimize recall bias about their experience. Potential participants were informed of the study purpose and asked if they would be interested in participating. We approached every 3^rd^ group of day hikers and every trail running group. Those who met the inclusion criteria (≥18 years of age, literate in English, returning from a hike or run that was <24 hours in duration and >2 miles) and were willing to participate were invited to complete the anonymous survey on a tablet. The first section of the survey included information required for informed consent. Given the wilderness context of the study, the use of tablets to administer the survey digitally, and verbal consent procedures, participants did not receive a copy of the consent form. All participants hiked or ran in wilderness designated areas. If hikers/runners were in a group, only one person per group was eligible to take the survey. For groups with more than 1 person, the person with the next birthday was asked to take the survey to eliminate volunteer bias. The survey was developed and implemented in REDCap, and administered via tablets in offline mode. Survey responses were uploaded to REDCap once internet access was available. The study was approved with an exempt determination by the Boston University Medical Campus Institutional Review Board.

The majority of survey questions were multiple choice or spectrum based; some were optional and open response ^12,27^. Hikers and trail runners self-selected their level of wilderness experience (none, little, some, experienced), days/years of wilderness experience, self-designated preparedness, equipment carried, health status, demographics, residence, hike completion status, altitude experience, and health problems or safety concerns encountered during the hike. Most survey questions were intended for the individual answering the survey, but some specified considering everyone in the group.

We are not aware of any standardized convention for determining day hiker/trail runner preparedness, or altitude preparedness. Consequently, we developed our own scale to assess each of these, utilizing previously published literature ^12,13,27^, wilderness educational information, and feedback from experienced outdoors enthusiasts. Participants were categorized as ‘wilderness prepared’ if they had at least 1L water, AND 7 out of 13 of the ‘10 essentials’, AND a map or equivalent, AND EITHER two rescue devices OR 8 out of 15 recommended first aid items. The rationale for this scale is that prepared visitors should have adequate hydration, carry the ‘essentials’, be able to orient themselves, and be able to signal for help or be equipped for basic first aid in the event of an injury or illness ^28^. We included 13 items on our list of ‘10 essentials’ because different sources include different ‘essentials’, and while the majority overlap, some participants may reasonably opt to carry different items depending on their source of information^29–32^.

Similarly, we utilized a scale to determine if participants were prepared for high altitude. Participants who scored three or more points were categorized as “altitude prepared”. One point was awarded for each of the following: having an altimeter or equivalent (topographic map + compass or a GPS device), taking or considering pharmacologic prophylaxis for altitude illness, self-reporting a safe ascent plan, and knowing at least two of the major symptoms of AMS (headache, emesis, fatigue, difficulty sleeping, tachycardia, irritability, dyspnea, and dizziness)^10^. Synonyms for certain AMS symptoms were accepted as correct. This scale is identical to that used in a previous survey study conducted in RMNP in 2018 ^12^.

Data were stored and analyzed in REDCap, Microsoft Excel 2016 (Microsoft Corp, Redmond, Washington) and R (version 4.2.3)^33^. Data cleaning included removing extreme values likely to be typos (e.g. age -35 removed). We performed descriptive statistics and multivariate logistic regression analysis to quantify the association between experience and preparedness, adjusting for activity, trip time, sex, age, and group size. Continuous variables were tested for statistical significance using an independent 2-sample T-test and categorical variables were tested with Pearson’s chi-squared test for independence. P ≤ 0.05 was used to determine statistical significance. Based on *a priori* power calculation, we planned to enroll at least 515 participants, as this number was estimated to provide sufficient sample size to find a 15% absolute difference in the proportions of groups that are ‘prepared’ using chi-squared single-variable analysis, with extra statistical power for multivariate logistic analysis. We express our findings of the regression analysis as odds ratio (OR) with 95% confidence intervals (CI).

## Results

We invited 801 groups or individuals (712 day hikers and 89 trail runners) to participate in the study. Of these, 82.3% (n=586) of day hikers and 76.4% (n=68) of trail runners consented to participate (median age 38, range 18–82, IQR 28–52 y). Approximately half of the respondents were women (50.1%), and the most common state of residence was Colorado (47.3%). Respondents represented a total of 1,714 individuals from 44 US states and 15 countries.

### Differences between groups

Compared to day hikers, trail runners were more likely to be male, younger, and travel in smaller groups (Table 1). Group sizes ranged from 1 to 20 for day hikers (median 2) and from 1 to 5 (median 1) for trail runners. Day hikers reported significantly fewer days of wilderness experience per year (30.1 ± 37.3 vs. 92.7 ± 86.1, mean ± SD) compared to trail runners. Trail runners also tended to have more days of wilderness experience per year, self-reported higher wilderness experience levels, and were more likely to have previously experienced a wilderness injury/illness. Trail runners reported going “significantly off trail” at much higher rates than day hikers (26.9% vs. 3.3%). Trail runners were more frequently only recreating in the wilderness for 1 day, whereas day hikers reported recreating in the wilderness for more consecutive days. Trail runners self-reported higher levels of physical fitness than day hikers; few (34) participants in either group self-reported low fitness. More than 90% of participants reported completing their planned hike or run.

**Table 1:** Characteristics of day hikers and trail runners in Rocky Mountain National Park (RMNP) in Summer 2024.

<sup>2</sup> Design-based KruskalWallis test; Pearson's X<sup>2</sup>: Rao & Scott adjustment.
|  | **Overall**<br>N = 654 | **Day Hiker**<br>N = 586 | **Trail Runner**<br>N = 68 | **p-value<sup>2</sup>** |
| --- | --- | --- | --- | --- |
| **Age (y), median**<br>(p25, p75) | 38 (28, 52) | 39 (28, 53) | 32 (28, 45) | 0.019 |
| **Age category**<br>(y), n (%) |  |  |  | 0.108 |
| 18-29 | 201 (31%) | 176 (30%) | 25 (37%) |  |
| 30-39 | 142 (22%) | 122 (21%) | 20 (30%) |  |
| 40-49 | 117 (18%) | 106 (18%) | 11 (16%) |  |
| 50-59 | 97 (15%) | 91 (16%) | 6 (9.0%) |  |
| 60+ | 94 (14%) | 89 (15%) | 5 (7.5%) |  |
| **Group size,**<br>median (p25,<br>p75) | 2.0 (2.0, 3.0) | 2.0 (2.0, 3.0) | 1.0 (1.0, 2.0) | <0.001 |
| **Sex, n (%)** |  |  |  | 0.034 |
| Male | 316 (48%) | 273 (47%) | 43 (63%) |  |
| Female | 324 (50%) | 300 (51%) | 24 (35%) |  |
| Other/Unknown§ | 14 (2.1%) | 13 (2.2%) | 1 (1.5%) |  |
| **Wilderness**<br>**experience level,**<br>n (%) |  |  |  | <0.001 |
| None/Little | 115 (18%) | 113 (20%) | 2 (3.1%) |  |
| Some/Experienced | 522 (82%) | 459 (80%) | 63 (97%) |  |
| **Experienced**<br>**safety concerns**<br>**or ‘close calls’**<br>**today, n (%)** | 44 (6.8%) | 35 (6.0%) | 9 (13%) | 0.023 |
| **Informed third**<br>**party of plans**<br>**and when to**<br>**expect them**<br>**back, n (%)** | 479 (74%) | 423 (73%) | 56 (84%) | 0.060 |
| **Wilderness**<br>**prepared, n (%)**<br>† | 109 (17%) | 92 (16%) | 17 (25%) | 0.052 |
| **Altitude**<br>**prepared, n (%)**<br>‡ | 260 (41%) | 228 (40%) | 32 (49%) | 0.166 |
| **Consecutive**<br>**days planning to**<br>**be in the**<br>**wilderness, n**<br>**(%)** |  |  |  | <0.001 |
| 1 day (today<br>only) | 291 (46%) | 247 (43%) | 44 (68%) |  |
| 2-3 days | 144 (23%) | 137 (24%) | 7 (11%) |  |
| 4-6 days | 111 (17%) | 107 (19%) | 4 (6.2%) |  |
| 7+ days | 89 (14%) | 79 (14%) | 10 (15%) |  |
| **Number of essential items carried, median (p25, p75)** | 6.0 (3.0, 8.0) | 6.0 (3.0, 8.0) | 6.0 (3.0, 8.0) | 0.806 |
| **Number of first aid items carried by group, median (p25, p75)** | 2.0 (1.0, 4.0) | 2.0 (1.0, 4.0) | 1.0 (0.0, 3.0) | 0.002 |

**Table 2:**
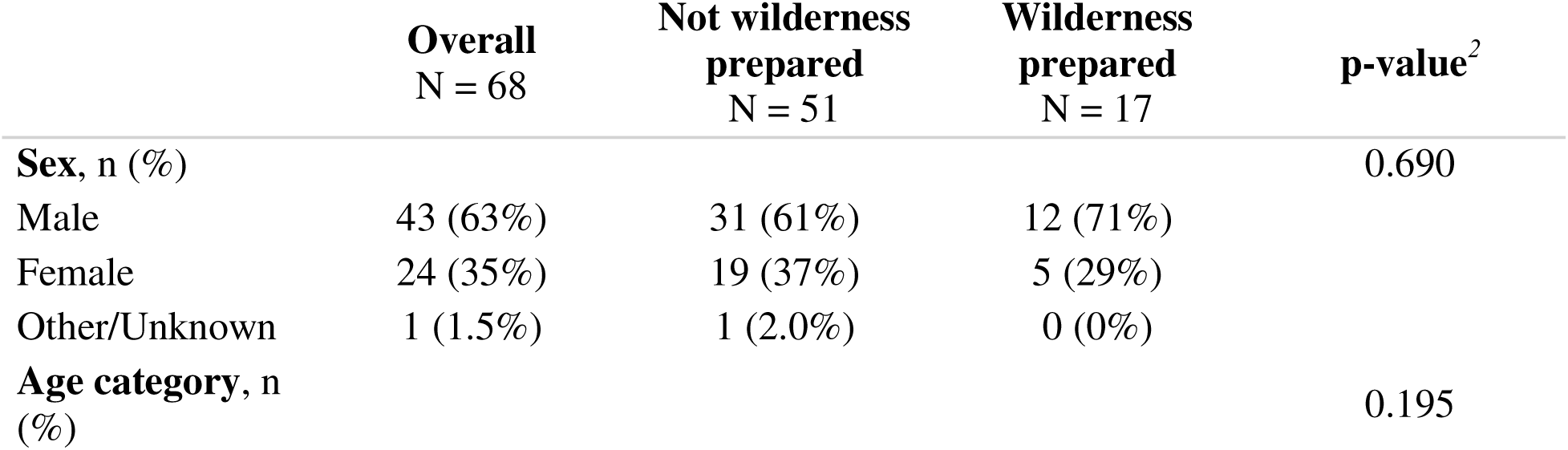
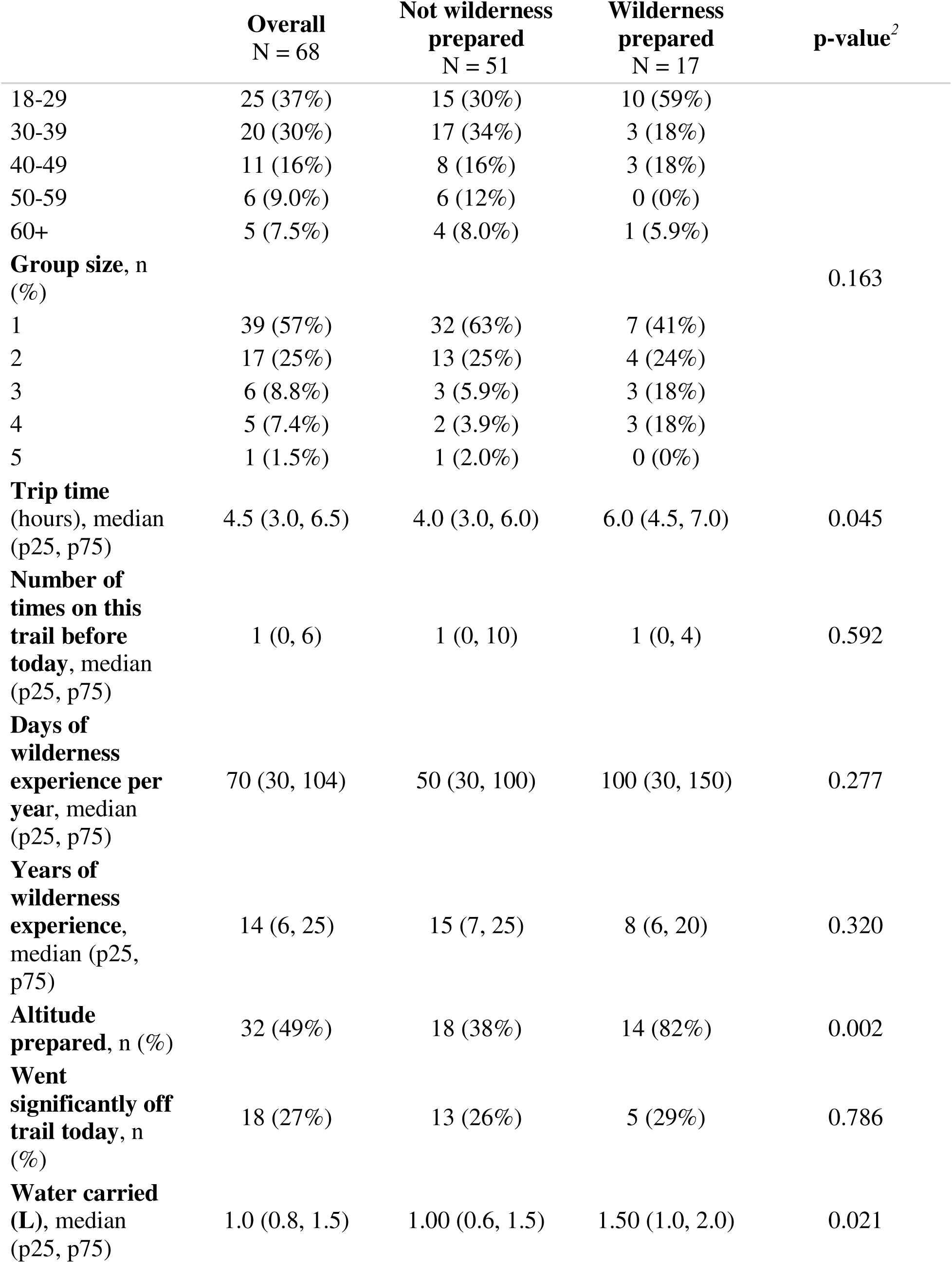
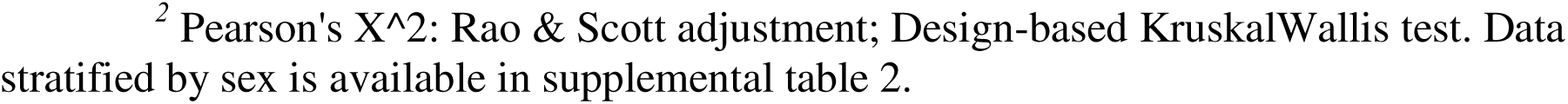
Characteristics of trail runners by level of wilderness preparedness.

|  | Overall<br>N = 68 | Not wilderness<br>prepared<br>N = 51 | Wilderness<br>prepared<br>N = 17 | p-value<sup>2</sup> |
| --- | --- | --- | --- | --- |
| **Sex, n (%)** |  |  |  | 0.690 |
| Male | 43 (63%) | 31 (61%) | 12 (71%) |  |
| Female | 24 (35%) | 19 (37%) | 5 (29%) |  |
| Other/Unknown | 1 (1.5%) | 1 (2.0%) | 0 (0%) |  |
| **Age category, n (%)** |  |  |  | 0.195 |

Table 2: <sup>2</sup> Pearson's X<sup>2</sup>: Rao & Scott adjustment; Design-based KruskalWallis test. Data stratified by sex is available in supplemental table 2.
|  | **Overall**<br>N = 68 | **Not wilderness**<br>**prepared**<br>N = 51 | **Wilderness**<br>**prepared**<br>N = 17 | **p-value<sup>2</sup>** |
| --- | --- | --- | --- | --- |
| 18-29 | 25 (37%) | 15 (30%) | 10 (59%) |  |
| 30-39 | 20 (30%) | 17 (34%) | 3 (18%) |  |
| 40-49 | 11 (16%) | 8 (16%) | 3 (18%) |  |
| 50-59 | 6 (9.0%) | 6 (12%) | 0 (0%) |  |
| 60+ | 5 (7.5%) | 4 (8.0%) | 1 (5.9%) |  |
| **Group size, n**<br>(%) |  |  |  | 0.163 |
| 1 | 39 (57%) | 32 (63%) | 7 (41%) |  |
| 2 | 17 (25%) | 13 (25%) | 4 (24%) |  |
| 3 | 6 (8.8%) | 3 (5.9%) | 3 (18%) |  |
| 4 | 5 (7.4%) | 2 (3.9%) | 3 (18%) |  |
| 5 | 1 (1.5%) | 1 (2.0%) | 0 (0%) |  |
| **Trip time**<br>(hours), median<br>(p25, p75) | 4.5 (3.0, 6.5) | 4.0 (3.0, 6.0) | 6.0 (4.5, 7.0) | 0.045 |
| **Number of**<br>**times on this**<br>**trail before**<br>**today, median**<br>(p25, p75) | 1 (0, 6) | 1 (0, 10) | 1 (0, 4) | 0.592 |
| **Days of**<br>**wilderness**<br>**experience per**<br>**year, median**<br>(p25, p75) | 70 (30, 104) | 50 (30, 100) | 100 (30, 150) | 0.277 |
| **Years of**<br>**wilderness**<br>**experience,**<br>**median (p25,**<br>**p75)** | 14 (6, 25) | 15 (7, 25) | 8 (6, 20) | 0.320 |
| **Altitude**<br>**prepared, n (%)** | 32 (49%) | 18 (38%) | 14 (82%) | 0.002 |
| **Went**<br>**significantly off**<br>**trail today, n**<br>(%) | 18 (27%) | 13 (26%) | 5 (29%) | 0.786 |
| **Water carried**<br>**(L), median**<br>(p25, p75) | 1.0 (0.8, 1.5) | 1.00 (0.6, 1.5) | 1.50 (1.0, 2.0) | 0.021 |

The overall proportion of those who experienced injuries or illness on the day of the survey was low; 2.9% of hikers and 4.5% of runners. The most common injuries were ankle injuries, and the most common illness was AMS. Significantly more trail runners reported experiencing safety concerns or ‘close calls’ on the day they participated, compared to day hikers (13.4% of runners, 6% hikers, p=0.023). The most common of these close calls were weather related (e.g. being above tree line in a thunderstorm), followed by falls and near falls.

Nearly all day hikers (87.7%) and trail runners (88.2%) categorized themselves as ‘adequately prepared’ or higher; however, only a minority of day hikers (15.7%) and trail runners (25.0%) were classified as wilderness prepared using our measure. Less than half were categorized as altitude prepared. Fewer day hikers informed a third party of their plan for the day or when to expect them back, compared to trail runners (27.1% of hikers, 16.4% of runners). On average, day hiker groups reported carrying more first aid items than trail runners. Individuals from both groups reported carrying about the same number of essential items.

Extra food and extra clothing were the most commonly carried essential items among respondents (Figure 2). Day hikers were significantly more likely to carry extra water, a first aid kit, and a knife compared to trail runners. Trail runners were significantly more likely to carry a light source and water treatment method compared to day hikers.

**Figure 2:**
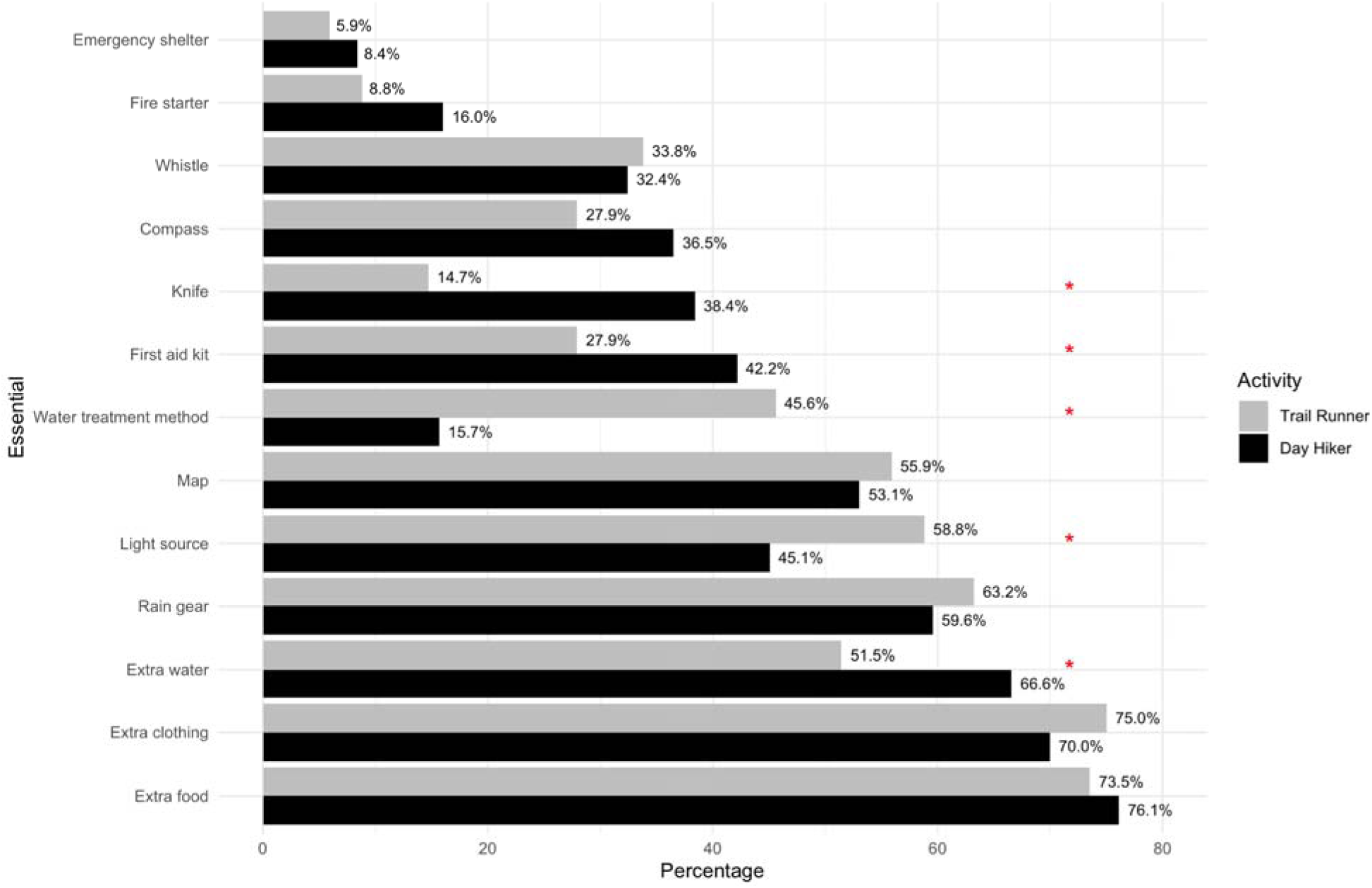
Percentage of individuals carrying each essential item by activity. Statistical significance marker: *p<0.05

Figure 3 shows the proportion of specific types of navigation and communication gear carried by a respondent or someone in their group. Nearly all participants/groups carried a cell phone. Trail runners were significantly more likely to carry a GPS, altimeter, two-way satellite messenger device, and personal locator beacon (PLB) compared to day hikers. Just over half of all groups carried a map during their wilderness day hike or trail run.

**Figure 3:**
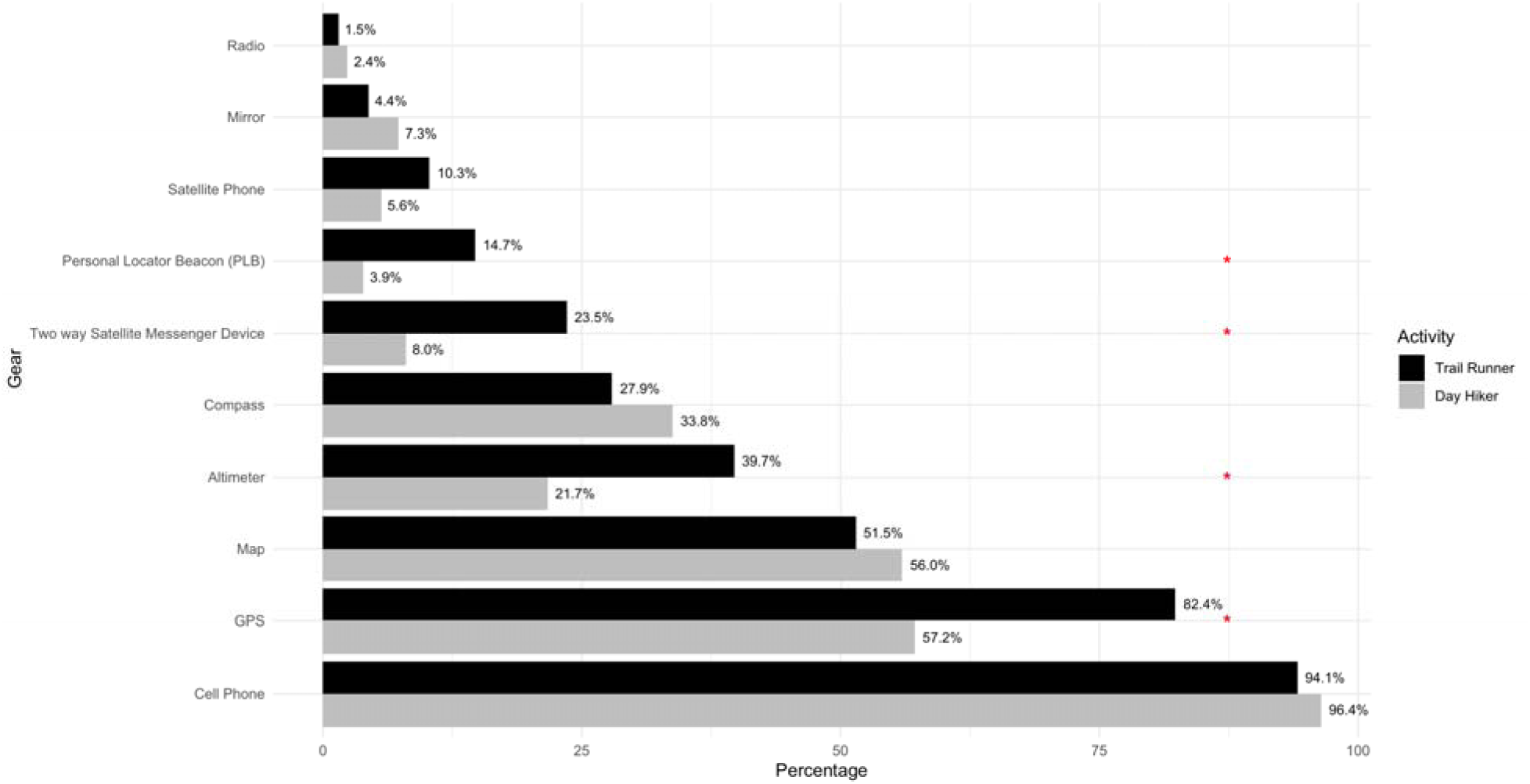
Percentage of people/groups carrying navigation and communication gear by activity. Statistical significance marker: *p<0.05; GPS, global positioning system.

Sunscreen was the most common first aid item carried for both groups. Day hikers were significantly more likely to carry sunscreen and bandage/tape compared to trail runners (Figure 4). Overall, the majority of groups did not carry most of the 15 first aid items asked about in the survey.

**Figure 4:**
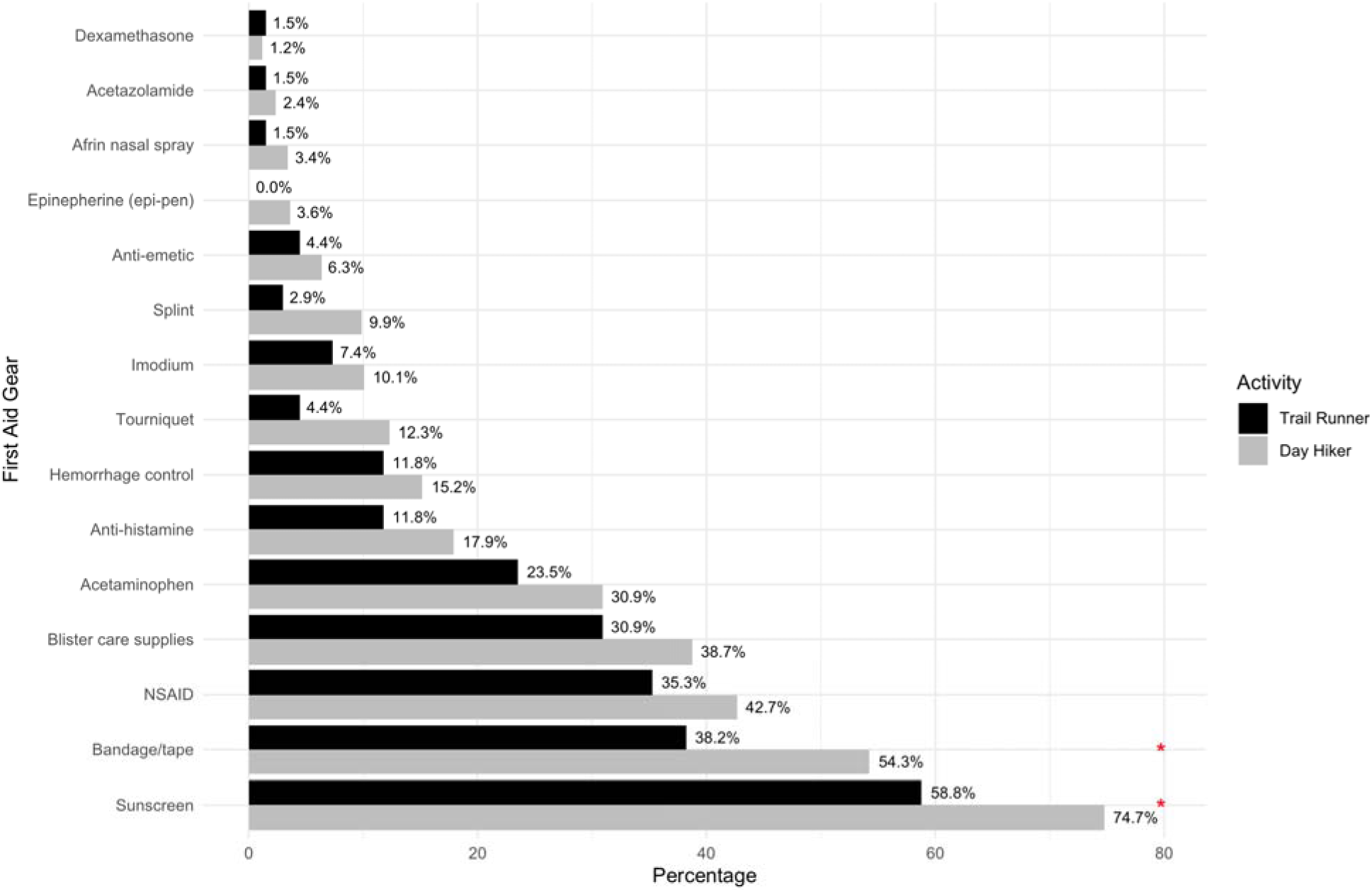
Percentage of people/groups carrying each first aid item by activity. Statistical significance marker: *p<0.05; NSAID, nonsteroidal anti-inflammatory drug.

### Trail runner characteristics

We encountered relatively few trail runners while surveying, 142 individuals (compared to 5,604 individual day hikers and 270 individuals doing other activities). Among trail runners, those who reported scrambling during their run were significantly more likely to report having gone “significantly off trail” (8.7% vs. 36.4%). Male trail runners were significantly more likely to report going “significantly off trail” during their run, compared to female participants (Supplemental Table 1). Those who reported scrambling also rated their run as more difficult, went on longer runs, carried more first aid and essential items, and were more likely to live in Colorado (Supplemental Table 2).

### Wilderness preparedness and experience

Our survey asked several different questions related to experience. We then fit a model to examine overall experience related predictors to wilderness preparedness and found key predictors related to experience: trip length, days spent in wilderness settings per year, higher self-reported experience level, and prior wilderness injury history all significantly increase the odds someone was wilderness prepared (Table 3). Self-reporting being "Some/Experienced" vs. "None/Little" was associated with more than double the odds of preparedness, each additional day spent in the wilderness per year was associated with slightly higher odds (∼0.5% per day) of preparedness, each additional hour of trip time was associated with higher odds of preparedness by about 21% per hour, having experienced a prior wilderness injury/illness was associated with higher odds of preparedness by ∼80%, possibly reflecting increased caution or experience following injury. The number of years of wilderness experience reported did not significantly change the odds of being wilderness prepared, nor did the number of times a person had been on that trail before.

**Table 3:** Predictors of wilderness preparedness.

|  | **OR** | **95% CI** | **p-value** |
| --- | --- | --- | --- |
| **Sex** |  |  |  |
| Male | — | — |  |
| Female | 1.01 | 0.64, 1.61 | >0.9 |
| Other/Unknown | 0.49 | 0.05, 4.55 | 0.5 |
| **Age (y)** | 1.01 | 0.99, 1.03 | 0.3 |
| **Group size** | 0.99 | 0.87, 1.12 | 0.9 |
| **Self-reported wilderness experience level** |  |  |  |
| None/Little | — | — |  |
| Some/Experienced | 2.32 | 1.04, 5.16 | 0.040 |
| **Activity** |  |  |  |
| Day hiker | — | — |  |
| Trail runner | 1.03 | 0.48, 2.22 | >0.9 |
| **Trip time (hours)** | 1.21 | 1.09, 1.33 | <0.001 |

Table 3: Abbreviations: CI = Confidence Interval, OR = Odds Ratio
|  | OR | 95% CI | p-value |
| --- | --- | --- | --- |
| **Number of times on this trail before today** | 1.00 | 1.00, 1.00 | >0.9 |
| **Days of wilderness experience per year** | 1.01 | 1.00, 1.01 | 0.015 |
| **Years of wilderness experience** | 0.98 | 0.96, 1.00 | 0.10 |
| **Experienced prior wilderness injury/illness** | 1.80 | 1.09, 2.98 | 0.022 |

Those who were wilderness prepared were significantly more likely to have told a third party of their plans, to have been at high altitude before, to be altitude prepared, on trail for more hours, and live in Colorado. There was no significant association between wilderness preparedness and age, sex, group size, talking to NPS staff or reading NPS materials, amount of water carried, trip completion status, or injury/illness on the day of the survey.

## Discussion

### Differences between groups

In this survey of 654 recreational wilderness day hikers and trail runners at RMNP, we found significant differences between day hikers and trail runners – runners tended to be younger, were more often male, experienced more safety concerns on trail, and tended to be more experienced overall. ^20,34^ More frequently, trail runners traveled alone and reported going “significantly off trail”, likely increasing their risk if injured. While most day hikers and trail runners informed a third party of their plans and when to expect them back, a substantial minority did not. Additionally, both day-use visitor groups overestimated their level of wilderness and altitude preparedness.

Our findings differ from prior literature in several important ways. We found that about half of day hikers were female, while a majority (63%) of trail runners were male, while previous studies of similar hiking populations in other outdoor areas and times generally found ∼60% male ^11,17,35^. This finding may indicate that the gender distribution of day hikers may have changed post-pandemic. Our study found no difference in or association with sex on wilderness preparedness, differing from Yue et al.’s findings ^12^. The number of essentials carried in our study was generally lower than in other studies in New Hampshire (NH) and Colorado, though this is likely explained by the exclusion of overnight backpackers from our study and inclusion in others ^13,27^. The number of essential items carried in our study (mean = 5.6, ± 3.05, median 6) was slightly higher than that found in 2016 by Daniel et al. (mean = 5, ± 2, median 5) in NH day hikers ^36^. Daniel et al. reported that the most commonly needed ’10 essentials’ were the same top 4 (extra food, clothing, water, rain gear) carried in our sample, though our study found a low proportion carrying a first aid kit – a key piece of gear to prepare for common adverse events. In our study, 11% of all participants reported carrying between 10-13 of the ’10 essentials’ items, while 27.2% of participants reported carrying >7 of the essentials. This is lower than what Mason et al. (2011) found, in which 40.6% carried >7 and 17.8% all 10 essentials in NH ^27^. This may be due to small sample size (n=199) and very different areas (NH) and years (2011).

### Trail runner characteristics

This study is the first of its kind to evaluate wilderness preparedness among trail runners compared to general wilderness preparedness. Trail runners were found to have distinct characteristics and patterns. Trail runners reported experiencing safety concerns or ‘close calls’ in higher proportions than day hikers. This may be due to their higher speed and resultant reaction time differences, or may be due to other factors. Notably, a high percentage of trail runners went “significantly off trail” (26.9%), especially those who reported scrambling. These trail runners are typically more difficult to find if a search is initiated. In recent years, multiple trail runners have gone missing or died while off trail or scrambling, and SAR teams have had difficulty finding or reaching them ^20,34,37^. These findings, taken together, illustrate the importance of trail runners prioritizing measures to ensure their safety, even more so than for day hikers. Our results confirm findings from a recent study of Brazilian trail runners that reported high rates of injuries (36%) in the past 12 months ^17^. Just over 40% of runners in our study reported previously experiencing a wilderness injury or illness. Though ‘only’ 4.5% of runners reported experiencing an injury or illness on the day of surveying, when extrapolated to the entire population this is a large number of people experiencing injuries and illnesses while trail running.

### Wilderness preparedness and experience

We identified multiple measures of experience that were associated with wilderness preparedness. Our findings suggest that visitors who spend fewer days in the wilderness per year, who are on shorter trips, and who identify as less experienced were less wilderness prepared.

Consequently, preventative search and rescue (PSAR) efforts for preparedness may be most effective when targeting these groups. Those who have previously experienced a wilderness injury or illness seem to learn from this experience, and were significantly more likely to be prepared in our sample. Our findings further suggest that PSAR should not assume that visitors with more years of wilderness experience, age, or familiarity with the trail are more wilderness prepared. Previous research shows mixed evidence on the impact of age on wilderness preparedness, but generally agrees with our finding that experience increases preparedness ^11–13^.

### Overall

Wilderness day hikers and trail runners generally must be in good physical shape, and it is unsurprising that most of the participants meet this requirement. Nearly half of the study participants are somewhat local (live in Colorado), and are therefore likely at least partly acclimated to altitudes experienced in the park. However, a majority of the study population does not live at altitude, and cannot let youth or fitness substitute for their own preparedness.

RMNP, like most wilderness areas, has generally poor cellular coverage. For this reason, it’s important for wilderness users to carry an alternative method to signal for help in case something goes wrong. While nearly every group/individual carried a cell phone, it should not currently be relied upon as a way to signal for help. A higher number of hikers and runners than expected carried alternative methods such as 2 way satellite messaging devices, satellite phones, and PLBs. However, these numbers were still relatively small; less than a quarter of hiking groups and less than half of running groups had an alternative way to call for help beyond a cell phone. This may change in the coming years, as phone manufacturers are increasingly adding satellite SOS and 2 way satellite messaging to new cell phones. In 10 years, carrying an extra battery/cord to charge a satellite-capable cell phone may be just as effective as carrying one of these dedicated devices today. Currently, only a few cell phones are able to connect to satellites, but this market is expected to expand rapidly, and may increase both dependence on smartphones and wilderness connectivity ^38^. This has not yet been studied in a systematic way, but may have major implications in the near future. Future studies should examine the impact of these phones, once they are commonplace.

As other groups have identified, just carrying 10 items from a list does not solely determine if one is ready to safely recreate in the wilderness ^36^. We categorized participants as prepared mainly based off of equipment carried, however, physical and mental preparation are also likely just as important as gear carried. Another consideration is that for a short hike, on a high traffic trail, certain items will be more useful or have a higher likelihood of use than others. Each wilderness user should be aware of the physical, mental, and equipment recommendations and prepare for each excursion accordingly. Wilderness users should ensure they know how to use all items they decide to carry.

### Strengths and limitations

Our study had several limitations. The items included on ’10 essentials’ lists vary widely, and our list of 13 items may not have included all items included on whichever list any individual may have consulted before packing ^32^. Additionally, some items on our list may vary by person (e.g. what they consider to constitute ‘extra clothing’). Trail runners declined to participate at a higher rate than day hikers, and fewer were recruited than planned. The most common reasons given for declining to participate in this study were a lack of time and unwillingness to stop. A handful of potential participants declined to participate due to dehydration/lack of water, hunger, fatigue, or weather. Potential subjects who declined to participate may represent a possible source of selection bias. Additionally, day hikers and trail runners who visit RMNP between September and May may represent a different population than our study cohort. Preparedness is difficult to assess with a written survey, and the instruments used here have not been validated, however our results are generally consistent with related literature. As with all surveys, there is variability in individual effort, however the authors believe that most participants put ample effort into answering. Due to low participant volume, no data collection occurred after dark, but this may have caused us to unintentionally exclude some of the least or best prepared hikers or runners. Lastly, our survey population only included individuals who were 18 years or older, so Scouts and Junior Rangers, who may be more well prepared, were excluded.

Notable strengths of this study include that this was the first of its kind to characterize wilderness trail runners and their preparedness. Data collection occurred in the field as participants were returning, so recall bias was minimized and data are high quality. Nearly all other wilderness survey studies have a limitation in which they may have missed participants who required SAR. In our study, no participants reported injuries severe enough that they planned to go to a clinic or hospital for treatment, but 1 SAR incident occurred on the same trail during the survey. The incident required assistance from the field researcher (JTL) and data collection was halted for the day. The individual requiring assistance was not asked to participate in the study; they were a day hiker requiring hospital-level treatment. Other SAR incidents occurred in other parts of the park or at different times from data collection, but did not impact participation, data collection or findings.

Notwithstanding this study’s limitations, our findings have implications for improving visitor preparedness and safety, by providing insight into wilderness day users. While the majority of both hikers and trail runners informed someone of where they were going and when to expect them back, increasing PSAR efforts to get those percentages closer to 100% would have a low cost/effort and high reward. When SAR is required, time is critical. A timely alert to an overdue person and accurate information about their planned whereabouts is likely to save lives ^39^. Our finding that users who have previously experienced a wilderness injury/illness are more likely to be prepared illustrates that past experience is more predictive of preparedness than many other attributes. With only 15.7% of hikers and 25% of trail runners categorized as wilderness prepared, it is clear that ample opportunity for PSAR exists. PSAR seeks to educate wilderness visitors and reduce the number of visitors requiring SAR assistance. Visitors who are more prepared may be better able to quickly and accurately request help, self-rescue, or not need SAR at all. While RMNP already has PSAR efforts in place, this study illustrates where current strategies may require refinement or additional resources. However, RMNP is just one of many national parks, and itself is surrounded by national and state forests. Millions of wilderness hours are spent in similar environments, and it’s important that day hikers and trail runners develop and utilize skills and equipment whenever they are in wilderness settings. RMNP and other parks may be ideal locations for teaching and equipping people to be safe no matter where or when they recreate in nature. The results from this study may help inform better direct messaging during PSAR efforts to those who are least prepared or at elevated risk.

## Conclusion

Wilderness day hiking and trail running are demanding activities, especially at the high altitude and rapidly changing weather of RMNP. The results of this study strengthens our understanding of the wilderness trail runners and day hikers. Wilderness travel has certain inherent risk, both this study and prior work illustrate that injuries and illnesses can happen to anyone in the wilderness ^35,40,41^. Overall wilderness preparedness was highly variable but generally low – possibly due to limited experience and high cost of equipment. There are a variety of factors that lead to SARs, but increased education and preparedness have been shown to decrease SAR missions. Aiming educational interventions at those at highest risk or lowest preparedness may help increase preparedness and reduce SAR events in the future. Should an incident occur on the trail, the success of safely leaving the wilderness depends in large part on the equipment and knowledge of those involved.

## Data Availability

The datasets generated during and/or analyzed during the current study are available from the corresponding author on reasonable request.

## Acknowledgements

We thank Mike Lukens and Paige Lambert, of the National Park Service, for their support. We also thank the Rocky Mountain Conservancy, Rocky Mountain National Park, and the Center for Climate and Health.

## Author Contributions

Conceptualization [JTL TNW], Data curation [JTL], Formal Analysis [JTL], Funding Acquisition [JTL, GAW], Investigation [JTL], Methodology [JTL GAW DHH TNW], Project administration [JTL], Resources [JTL GAW DHH], Software [JTL], Supervision [JTL GAW DHH], Validation [JTL], Visualization [JTL], Writing – original draft [JTL], Writing – review and editing [JTL GAW DHH TNW]

## Statements and Declarations

### Ethical Considerations

The study was approved under exempt determination by the Boston University Medical Campus Institutional Review Board (IRB Number: H-44879) on May 5, 2024. We obtained the necessary permit to conduct research in RMNP (Permit Number: ROMO-2024-SCI-0012) on May 15, 2024.

### Consent to Participate

Participants were verbally consented for participation in this study. Written consent was waived by the BUMC IRB. No personally identifiable data were collected.

### Consent for Publication

Not applicable.

## Supplement

Survey Questions

**Supplemental Figure 1:**
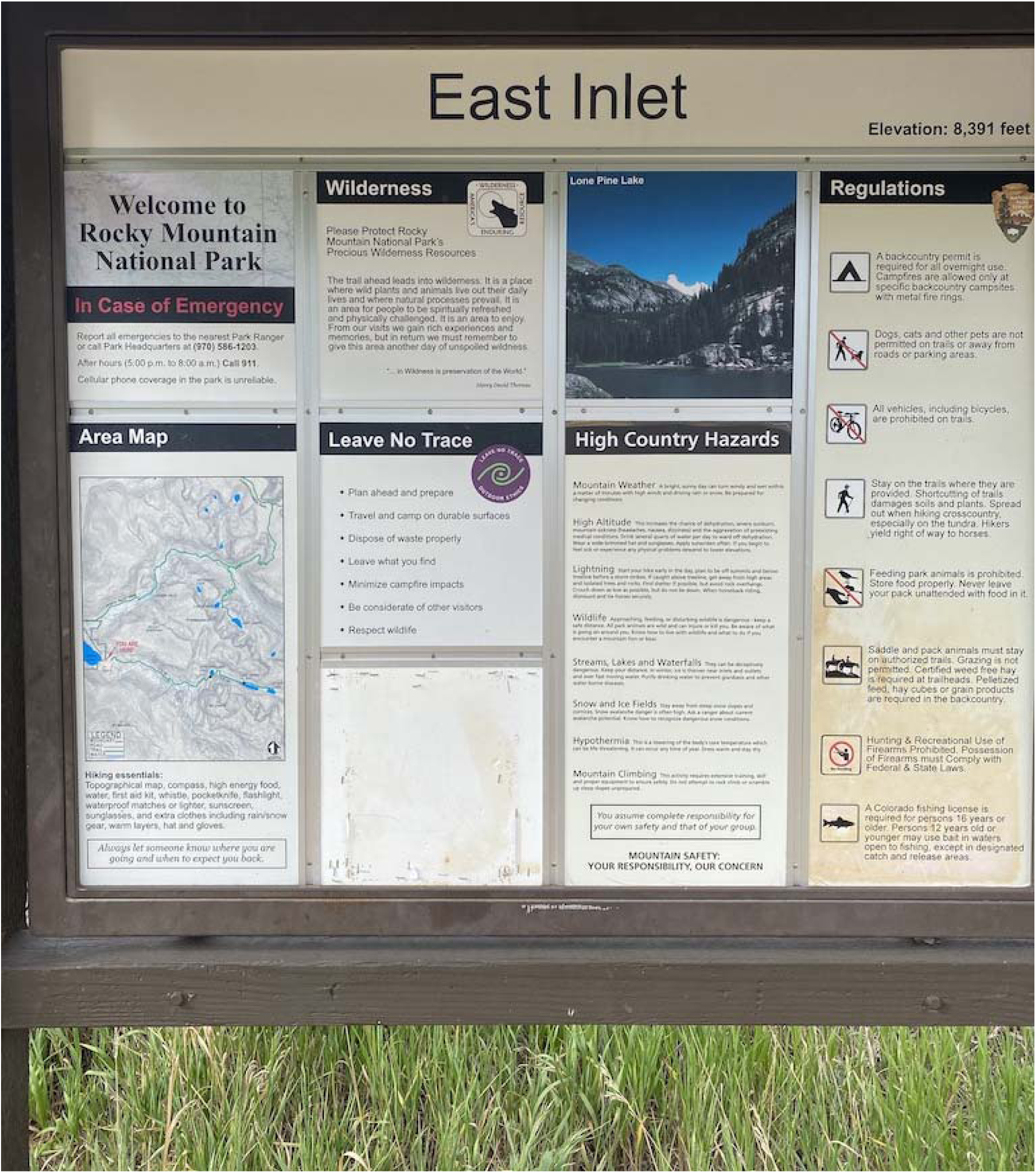
NPS sign at one of the trailheads in RMNP.

**Supplementary Table 1:** Characteristics of trail runners, stratified by sex.

|  | **Female**<br>N = 24<sup>1</sup> | **Male**<br>N = 43<sup>1</sup> | **p-value**<sup>2</sup> |
| --- | --- | --- | --- |
| **Age category (y), n (%)** |  |  | 0.454 |
| 18-29 | 12 (52%) | 13 (30%) |  |
| 30-39 | 5 (22%) | 14 (33%) |  |
| 40-49 | 3 (13%) | 8 (19%) |  |
| 50-59 | 1 (4.3%) | 5 (12%) |  |
| 60+ | 2 (8.7%) | 3 (7.0%) |  |
| **Group size, n (%)** |  |  | 0.509 |
| 1 | 11 (46%) | 27 (63%) |  |
| 2 | 7 (29%) | 10 (23%) |  |
| 3 | 3 (13%) | 3 (7.0%) |  |
| 4 | 2 (8.3%) | 3 (7.0%) |  |
| 5 | 1 (4.2%) | 0 (0%) |  |
| **Trip time (hours), median (p25, p75)** | 4.00 (3.00, 6.50) | 5.00 (3.00, 6.00) | 0.339 |
| **Number of times previously on this trail, median (p25, p75)** | 0 (0, 4) | 2 (0, 10) | 0.165 |
| **Days of wilderness experience per year, median (p25, p75)** | 80 (30, 150) | 52 (30, 100) | 0.692 |
| **Years of wilderness experience, median (p25, p75)** | 14 (6, 25) | 13 (7, 25) | 0.600 |
| **Altitude prepared, n (%)** | 11 (50%) | 21 (50%) | >0.9 |
| **Wilderness prepared, n (%)** | 5 (21%) | 12 (28%) | 0.529 |
| **Went significantly off trail today, n (%)** | 1 (4.3%) | 17 (40%) | 0.003 |
| **Water carried (L), median (p25, p75)** | 1.00 (1.00, 1.50) | 1.00 (0.50, 1.50) | 0.574 |
| **Number of essentials, median (p25, p75)** | 5.0 (2.0, 8.0) | 6.0 (3.0, 8.0) | 0.438 |

<sup>1</sup> 1 participant is excluded from this table due to not providing sex data
<sup>2</sup> Pearson's X<sup>2</sup>: Rao & Scott adjustment; Design-based KruskalWallis test

**Supplementary Table 2:** Characteristics of trail runners, stratified by scrambling.

|  | **Did not scramble**<br>N = 23 | **Scrambled**<br>N = 44 | **p-value<sup>2</sup>** |
| --- | --- | --- | --- |
| **Age**, median (p25, p75) | 33 (29, 45) | 32 (27, 42) | 0.292 |
| **Difficulty**, n (%) |  |  | 0.092 |
| Very easy | 2 (8.7%) | 0 (0%) |  |
| Easy | 6 (26%) | 6 (14%) |  |
| Medium | 11 (48%) | 24 (55%) |  |
| Hard | 4 (17%) | 14 (32%) |  |
| Very hard | 0 (0%) | 0 (0%) |  |
| **Group size**, n (%) |  |  | 0.374 |
| 1 | 11 (48%) | 28 (64%) |  |
| 2 | 8 (35%) | 9 (20%) |  |
| 3 | 1 (4.3%) | 4 (9.1%) |  |
| 4 | 2 (8.7%) | 3 (6.8%) |  |
| 5 | 1 (4.3%) | 0 (0%) |  |
| **Trip time** (hours), median (p25, p75) | 3.00 (2.00, 4.00) | 6.00 (3.75, 7.00) | <0.001 |
| **Number of times previously on this trail**, median (p25, p75) | 0 (0, 5) | 2 (0, 10) | 0.239 |
| **Altitude prepared**, n (%) | 8 (36%) | 24 (56%) | 0.145 |
| **Wilderness prepared**, n (%) | 4 (17%) | 13 (30%) | 0.285 |
| **Went significantly off trail today**, n (%) | 2 (8.7%) | 16 (36%) | 0.019 |
| **State of residence**, n (%) |  |  | 0.172 |
| Colorado | 15 (68%) | 35 (83%) |  |
| Elsewhere | 7 (32%) | 7 (17%) |  |
| **Essentials carried**, median (p25, p75) | 4.0 (2.0, 7.0) | 6.0 (4.0, 8.0) | 0.020 |

1 participant is excluded from this table due to not providing scrambling data
<sup>2</sup> Design-based KruskalWallis test; Pearson's X<sup>2</sup>: Rao & Scott adjustment

## References

1. Statista. Number of participants in trail running in the U.S. 2006-2017. Updated July 23, 2025. Accessed September 23, 2025, https://www.statista.com/statistics/191333/participants-in-trail-running-in-the-us-since-2006/

2. Strava. Strava Year in Sport - The Trend Report. 2023. Accessed September 23, 2025. https://downloads.ctfassets.net/9olkiac82a1q/iU421iiO6Hky1MVvYThmz/9b6b0755d1698d8c875758c0e90da0f3/Strava-Global-Report-2023-en-US.pdf

3. Taff BD, Rice WL, Lawhon B, Newman P. Who Started, Stopped, and Continued Participating in Outdoor Recreation during the COVID-19 Pandemic in the United States? Results from a National Panel Study. Land. 2021;10(12)doi:10.3390/land10121396

4. Annual Park Recreation Visits (1915 - Last Calendar Year). 2025. Accessed August 3, 2025. https://irma.nps.gov/Stats/Reports/Park/ROMO

5. Miller-Rushing AJ, Athearn N, Blackford T, et al. COVID-19 pandemic impacts on conservation research, management, and public engagement in US national parks. Biological Conservation. 2021/05/01/ 2021;257:109038. doi:10.1016/j.biocon.2021.109038

6. Park Statistics. National Park Service. Updated July 29, 2024. Accessed September 23, 2025, https://www.nps.gov/romo/learn/management/statistics.htm

7. Reynolds S, Beseler C, Stallones L. Preventative Search and Rescue for Rocky Mountain National Park (RMNP) Phase 1 Report. 2012. https://files.cfc.umt.edu/cesu/NPS/CSU/2011/11Reynolds_ROMO_searchrescue_fnl_rpt.pdf

8. Search and Rescue Needs Assessment 2019 (National Park Service) (2019).

9. Malcolm C, Hannah H, Pearce E. Effectiveness of Preventive Search and Rescue: Illness and Injury Prevention and Fiscal Impact. Wilderness & Environmental Medicine. 2014;25(3):355–356. doi:10.1016/j.wem.2014.01.014

10. Luks AM, Auerbach PS, Freer L, et al. Wilderness Medical Society Clinical Practice Guidelines for the Prevention and Treatment of Acute Altitude Illness: 2019 Update. Wilderness & Environmental Medicine. 2019/12/01 2019;30(4_suppl):S3-S18. doi:10.1016/j.wem.2019.04.006

11. Daniel NJ, Patel SB, St. Marie P, Schoenfeld EM. Assessment of “10 essential” preparedness among day-hikers on Mount Monadnock. The American Journal of Emergency Medicine. 2020/02/01/ 2020;38(2):401-403. doi:10.1016/j.ajem.2019.158382

12. Yue MDT, Spivey DW, Gingold DB, Sward DG. The Effect of Wilderness and Medical Training on Injury and Altitude Preparedness Among Backcountry Hikers in Rocky Mountain National Park. World journal of emergency medicine. 2018;9(3):172–177. doi:10.5847/wjem.j.1920-8642.2018.03.002

13. Brandenburg WE, Davis CB. Medical Knowledge and Preparedness of Climbers on Colorado’s 14,000-Foot Peaks. Wilderness & Environmental Medicine. 2016;27(1):62–68. doi:10.1016/j.wem.2015.11.009

14. Spano SJ, Hile AG, Jain R, Stalcup PR. The Epidemiology and Medical Morbidity of Long-Distance Backpackers on the John Muir Trail in the Sierra Nevada. Wilderness & Environmental Medicine. 2018/06/01 2018;29(2):203-210. doi:10.1016/j.wem.2018.02.006

15. Viljoen C, Janse van Rensburg DC, van Mechelen W, Verhagen E, Korkie E, Botha T. Development of a trail running injury screening instrument: A multiple methods approach. Physical Therapy in Sport. 2022/07/01/ 2022;56:60-75. doi:10.1016/j.ptsp.2022.06.003

16. Scheer V, Basset P, Giovanelli N, Vernillo G, Millet GP, Costa RJS. Defining Off-road Running: A Position Statement from the Ultra Sports Science Foundation. International journal of sports medicine. May 2020;41(5):275–284. doi:10.1055/a-1096-0980

17. Rizzo F, Vallio CS, Hespanhol L. HealthyTrailsBR – The prevalence of running-related injuries and cramps, and the description of personal and running characteristics in Brazilian trail runners: a cross-sectional study. Brazilian Journal of Physical Therapy. 2024/09/01/ 2024;28(5):101117. doi:10.1016/j.bjpt.2024.101117

18. Smiley A, Ramos WD, Elliott LM, Wolter SA. Association between trail use and self-rated wellness and health. BMC Public Health. 2020;20:1–10. doi:10.1186/s12889-020-8273-0

19. Butler LT, Field DT, Golightly M, Masento NA, van Reekum CM. Effects of hydration status on cognitive performance and mood. British Journal of Nutrition. 2014;111(10):1841–1852. doi:10.1017/S0007114513004455

20. Blevins J. Two runners have gone missing in remote Colorado mountains, leading to push for more education. The Colorado Sun. October 21, 2022. Accessed September 23, 2025. https://coloradosun.com/2022/10/21/trail-runners-education-mountain-safety/

21. Viljoen CT, Janse van Rensburg DC, Verhagen E, et al. Epidemiology of Injury and Illness Among Trail Runners: A Systematic Review. Sports Medicine. 2021/05/01 2021;51(5):917-943. doi:10.1007/s40279-020-01418-1

22. Vernillo G, Savoldelli A, La Torre A, Skafidas S, Bortolan L, Schena F. Injury and illness rates during ultratrail running. International journal of sports medicine. 2016;37(07):565–569. doi:10.1055/s-0035-1569347

23. Beery T, Olsson MR, Vitestam M. Covid-19 and outdoor recreation management: Increased participation, connection to nature, and a look to climate adaptation. Journal of Outdoor Recreation and Tourism. 2021/12/01/ 2021;36:100457. doi:10.1016/j.jort.2021.100457

24. Repanshek K. Longs Peak Is A Popular, But Potentially Deadly, Rocky Mountain National Park Destination. National Parks Traveler November 28, 2018. Accessed August 6, 2025. https://www.nationalparkstraveler.org/2018/11/longs-peak-popular-potentially-deadly-rocky-mountain-national-park-destination-2

25. Blotkamp A, Boyd W, Eury D, Hollenhorst S. Rocky Mountain National Park Visitor Study. Vol. 2011. Summer 2010. 2011/121/107587. https://s3.wp.wsu.edu/uploads/sites/3019/docs/235.1_ROMO_rept.pdf

26. Graham R, Miller A, Monz C. Post-Experience Survey of Backcountry Anglers and Hikers in Rocky Mountain National Park. All Graduate Plan B and other Reports. 2021;1569

27. Mason RC, Suner S, Williams KA. An Analysis of Hiker Preparedness: A Survey of Hiker Habits in New Hampshire. Wilderness & Environmental Medicine. 2013/09/01/ 2013;24(3):221-227. doi:10.1016/j.wem.2013.02.002

28. Brandenburg WE, Locke BW. Mountain medical kits: epidemiology-based recommendations and analysis of medical supplies carried by mountain climbers in Colorado. Journal of Travel Medicine. 2017;24(2):taw088. doi:10.1093/jtm/taw088

29. Ten Essentials. National Park Service; 2023. Accessed September 23, 2025. https://www.nps.gov/articles/10essentials.htm

30. Regenold S. The Scout 10 essentials: Items every Scout needs in the outdoors. Scouting. April 1, 2013. Accessed September 23, 2025. https://scoutingmagazine.org/2013/02/the-10-essentials/

31. REI. The Ten Essentials. Accessed September 23, 2025, 2024. https://www.rei.com/learn/expert-advice/ten-essentials.html

32. DeLoughery TG, DeLoughery EP. 10, 14, 19 Essentials? Wilderness & Environmental Medicine. 2025;36(3)doi:10.1177/10806032251342116

33. *R: A language and environment for statistical computing.* R Foundation for Statistical Computing; 2025. https://www.R-project.org/

34. Viljoen C, da Cruz M, Matlala K, et al. Trail Running Safety: A Review of Serious Adverse Events Reported in Online News Articles. Wilderness & Environmental Medicine. 2025;36(3):416-433. doi:10.1177/10806032251338703

35. Pearce EA, Jelínková L, Fullerton L, et al. Observational Study of Grand Canyon Rim-to-Rim Day Hikers: Determining Behavior Patterns to Aid in Preventive Search and Rescue Efforts. Wilderness & Environmental Medicine. 2019/03/01 2019;30(1):4-11. doi:10.1016/j.wem.2018.08.001

36. Daniel NJ, Patel SB, St. Marie P, Schoenfeld EM. Rethinking hiker preparedness: Association of carrying “10 essentials” with adverse events and satisfaction among day-hikers. The American Journal of Emergency Medicine. 2021/11/01/ 2021;49:253-256. doi:10.1016/j.ajem.2021.06.017

37. Zhang C. Search for trail runner in Rocky Mountain National Park called off after two weeks. The Colorado Sun. Oct 10, 2023. Accessed September 23, 2025. https://coloradosun.com/2023/10/10/search-for-trail-runner-in-rocky-mountain-national-park-called-off-after-two-weeks/

38. Siddiqui A, Grush A. Android and iPhone satellite connectivity: What is it and what are your options right now? Android Authority. Updated October 9, 2024. Accessed September 23, 2025, https://www.androidauthority.com/smartphone-satellite-connectivity-3295162/

39. Adams AL, Schmidt TA, Newgard CD, et al. Search Is a Time-Critical Event: When Search and Rescue Missions May Become Futile. Wilderness & Environmental Medicine. 2007/06/01 2007;18(2):95-101. doi:10.1580/06-WEME-OR-035R1.1

40. Heggie TW, Heggie TM. Viewing Lava Safely: An Epidemiology of Hiker Injury and Illness in Hawaii Volcanoes National Park. Wilderness Environ Med. 2004/06/01/ 2004;15(2):77-81. doi:10.1580/1080-6032(2004)015[0077:VLSAEO]2.0.CO;2

41. Kogut KT, Rodewald LE. A field survey of the emergency preparedness of wilderness hikers. Journal of Wilderness Medicine. 1994/06/01/ 1994;5(2):171–178. doi:10.1580/0953-9859-5.2.171

